# Evaluating Diuretics in Normal Care (EVIDENCE): A feasibility report of a pilot cluster randomised trial of prescribing policy in primary care to compare the effectiveness of thiazide-type diuretics in hypertension

**DOI:** 10.1101/2021.08.19.21262289

**Authors:** Angela Flynn, Amy Rogers, Lewis McConnachie, Rebecca Barr, Robert WV Flynn, Isla S Mackenzie, Thomas M MacDonald, Alexander SF Doney

## Abstract

**Background:** Obtaining evidence on comparative effectiveness and safety of widely prescribed drugs in a timely and cost-effective way is a major challenge for healthcare systems. Here we describe the feasibility of the Evaluating Diuretics in Normal Care (EVIDENCE) study that compares a thiazide and thiazide-like diuretics for hypertension as an exemplar of a more general framework for efficient generation of such evidence.

In 2011, the UK NICE hypertension guideline included a recommendation that thiazide-like diuretics (such as indapamide) be used in preference to thiazide diuretics (such as bendroflumethiazide) for hypertension. There is sparse evidence backing this recommendation, and bendroflumethiazide remains widely used in the UK.

**Methods:** Patients prescribed indapamide or bendroflumethiazide regularly for hypertension were identified in participating General Practices. Allocation of a prescribing policy favouring one of these drugs was then randomly applied to the Practice and, where required to comply with the policy, repeat prescriptions switched by pharmacy staff. Patients were informed of the potential switch by letter and given the opportunity to opt-out. Practice adherence to the randomised policy was assessed by measuring the amount of policy drug prescribed as a proportion of total combined indapamide and bendroflumethiazide. Routinely collected hospitalization and death data in the NHS will be used to compare cardiovascular event rates between the two policies.

**Results:** This pilot recruited 30 primary care practices in five Scottish National Health Service (NHS) Boards. Fifteen practices were randomised to indapamide (2682 patients), and 15 to bendroflumethiazide (3437 patients); a study population of 6119 patients. Prior to randomisation, bendroflumethiazide was prescribed to 78% of patients prescribed either of these drugs. Only 1.6% of patients opted out of the proposed medication switch.

**Conclusion:** The pilot and subsequent recruitment confirms the methodology is scalable within NHS Scotland for a fully powered larger study, currently 102 GP practices (>12,700 patients) are participating in this study. It has the potential to efficiently produce externally valid comparative effectiveness data with minimal disruption to practice staff or patients. Streamlining this pragmatic trial approach, has demonstrated the feasibility of a random prescribing policy design framework that can be adapted to other therapeutic areas.

**Trial registration number:** ISRCTN 46635087; registered pre-results, 11/08/2017.

**Summary:** We report on a Chief Scientist Office for Scotland-funded pilot of the feasibility of the (Evaluating Diuretics in Usual Care) EVIDENCE study. This report will describe:

- Recruitment and policy randomisation of 30 GP practices across 5 NHS health board regions in Scotland.
- Acceptability of study implementation information provided to primary healthcare practitioners and patients.
- Recruitment rates, staffing, training, and funding requirement estimations to inform the full-sized project.
- How knowledge and practical experience gained has informed scaling of activities to realise a fully powered EVIDENCE study, including 250 practices.

*Key messages regarding feasibility:*

- What uncertainties existed regarding the feasibility? For widely prescribed medicines with similar mode of action and similar indications differences in effectiveness are likely to be quite small indicating the need for very large study sizes. Previous work has demonstrated that practices would be reluctant to take part in this kind of study if it involved any extra work within already limited practice capacity. Would NHS Primary Care leads be willing to endorse the study taking place in their region? In addition, the large size and geographically distributed nature of this study meant devising solutions for work to take place remotely or using pre-existing regional staff needed to be devised. There was also uncertainty about the overall acceptability for patients in having their medication changed for research purposes.
- What are the key feasibility findings? The experience from the pilot and subsequent successful expanded recruitment shows that the solutions we developed seemed to be acceptable and achievable both for general practice staff and the patients they care for. Obtaining endorsement from key stakeholders in NHS health boards improved recruitment success with the practices and sustained support for the study. Negotiations with pharmacy regional leads around workforce implications enabled us to approach practices with a range of solutions for study implementation. It was found that offering training for the EVIDENCE study along with general clinical trials training for regional Pharmacists was key to recruitment of pharmacy delegates on a large scale. We were able to develop the IT infrastructure around the study to allow remote delivery of both training and implementation providing a framework that could be delivered during the covid-19 pandemic. We found that concerns from patients constituted a very small minority indeed indicating overwhelming tacit support for the objectives of the study
- What are the implications of the feasibility findings for the design of the main study? The initial engagement with a range of healthcare providers offered support for the study in areas that may otherwise have become a potential barrier to success. We will use this approach in the main study. Many of the practical skills required to undertake the EVIDENCE study were also useful skills for pharmacy teams in their everyday practice so acceptance of the study training and implementation was higher. We will continue to progress the study using this methodology as this provided beneficial professional development for pharmacy staff and a platform for a future research ready workforce. Demonstrating the success of this approach we have currently recruited over 100 practices and have 29 pharmacy delegates across Scotland outside the core study team. This indicates that recruiting the target of 250 practices (approximately 50,000 individuals is entirely feasible.

## Background

### The need for comparative effectiveness research

The National Health Service (NHS) has a responsibility to ensure the medications it uses are effective and safe. For many common long-term clinical conditions, prescribers can choose from a range of medicines with similar modes of action, but there is very often a lack of comparative effectiveness data to guide this choice within classes. As more “me-too” medicines are continually being developed (1), addressing this widening knowledge gap in a timely and efficient manner is a growing challenge for healthcare systems (2, 3).

Guidelines, such as those produced by the National Institute for Health and Care Excellence (NICE), provide clinicians with a reliable source of information to guide the delivery of evidence-based healthcare(4). The recommendations within these guidelines are based, where possible, on evidence provided by randomised controlled trials (RCTs), widely considered the gold standard. The disadvantages of RCTs are that these are often expensive, slow, and poorly applicable to NHS practice (poor external validity) because they include only highly selected participants (5, 6). Difficulties in recruitment to RCTs is also a barrier to their successful completion (7). This is an important issue as the results of clinical trials need to be more generalisable to the population to which they apply, including the elderly and those with multimorbidity. Responding to these challenges and the additional complications of research participation during COVID-19 the INCLUDE project, commissioned by the UK National Institute for Health Research Clinical Research Network, has recently produced guidance on how to improve the inclusion of under-represented groups in clinical research. (8)

Evidence from observational research using routinely collected healthcare data can be less expensive and quicker to produce but, due to challenges of controlling for bias, this evidence is often of insufficient standard to change prescribing behaviour. New efficient ways of doing high quality, externally valid, comparative effectiveness research are therefore required. The EVIDENCE (EValuatIng DiurEtics in Normal Care) study methodology addressed these issues using a hybrid study design combining features of both interventional and observational study design.

### Rationale for EVIDENCE

Hypertension affects 1 in 4 adults and is predicted to affect over 1.5 billion people worldwide by 2025. (9). It is one of the most important preventable causes of premature cardiovascular morbidity and mortality, being responsible for 54% of strokes and 47% of ischaemic heart disease(10). There are several classes of medicines used to treat hypertension; each includes agents with similar modes of action but unique pharmacological properties which may be clinically relevant. However, there remains a lack of reliable comparative effectiveness and safety data to inform prescribing on the most effective agent within each class. For medicines used to treat very common conditions like hypertension, even quite small differences in effectiveness or safety could have large public health consequences. On the other hand, the demonstration of clinical equivalence generates reassurance to choose medication based on price or availability.

One class of treatment, thiazide-type diuretics, have been the cornerstone of hypertension management since the late 1950’s(11). This was reviewed in 2011, and again in 2019 with NICE stating (12, 13), “where a thiazide-like diuretic is indicated, chlorthalidone or indapamide should be chosen in preference to a conventional thiazide diuretic such as bendroflumethiazide or hydrochlorothiazide”. However, the guideline development group conceded that further evidence was required to justify this position. The guidelines were disputed (14). Neither hydrochlorothiazide nor chlorthalidone are widely available to prescribers in the NHS and, despite the recommendation, community dispensed prescribing data for Scotland in 2018/2019 indicated that 86% of thiazide-type diuretic prescriptions were for bendroflumethiazide and only 14% for indapamide(15). Reasons the recommendations have not been fully adopted probably relate to familiarity, differential drug costs, and an assumption by many prescribers that the two medicines are clinically equivalent. Indeed, a recent meta-analysis by our group, whilst highlighting the need for further research, demonstrated no difference in outcomes between indapamide and bendroflumethiazide (16).

The overall aim of the EVIDENCE study is to compare the effectiveness of a policy of prescribing indapamide with a policy of prescribing bendroflumethiazide in the management of hypertension in NHS Scotland. The study methodology is adaptable as a framework for generating comparative effectiveness data for other therapeutic areas within the NHS. For example, within-class comparisons of commonly used medications such as angiotensin-converting enzyme inhibitors (ACEI) could be made to ascertain which of the many generic ACEIs are the most effective in clinical practice.

Medication switches for safety, financial or supply reasons are a widely accepted part of routine care in the NHS. A comparison of prescribing policy has been judged in this case not to constitute a clinical trial of investigational medicinal products (CTIMP) by the Medicines and Healthcare Regulatory Authority (MHRA). In addition, ethical opinion was that individual patients do not require to be consented because this study is comparing a prescribing policy intervention implemented in the same way as would occur for any routine prescribing policy change.

We have previously considered the ethical aspects (17) and have surveyed public and healthcare professional opinions, and found broad support for a study of this design (18).

## Methods

The EVIDENCE methods and protocol have been previously described in detail (19). In this report, we focus on the practical application of the EVIDENCE protocol, as experienced during the pilot study.

EVIDENCE is a prospective, cluster-randomised, open-label, blinded end-point study conducted in the primary care setting. It aims to formally test the equivalence of a policy of prescribing indapamide versus a policy of prescribing bendroflumethiazide in preventing major adverse cardiovascular events.

The study protocol has been iteratively developed throughout the pilot after broad consultation with healthcare providers, patients, as well as a patient involvement group. A critical aim of the study design is to be minimally disruptive to primary healthcare delivery by making use of the prescribing management systems already used in Scotland for routine large-scale medicine switching.

Scottish NHS primary care General Practices, hereafter referred to as practices, are the unit of randomisation in this study. The intervention is the implementation of a practice-level policy of prescribing bendroflumethiazide versus a policy of prescribing indapamide when a thiazide-type diuretic is indicated for the management of hypertension. Practices wishing to take part must first agree that they are in clinical equipoise between prescribing these diuretics and agree to be randomised to one or other prescribing policy.

EVIDENCE has been approved by the East of Scotland Research Ethics Service (REC Number 17/ES/0016) and registered with ISRCTN (46635087).

### Recruiting Practices

During the pilot study, we approached a range of key primary care stakeholders to generate awareness of, and obtain approval and support for delivering, the EVIDENCE study across Scotland. These included Associate Directors of Primary Care and Directors of Pharmacy, leads for Area Drugs and Therapeutics Committees and regional Primary Care Clinical Leads. Presentations were delivered at various regional and national primary healthcare conferences to generate awareness and interest in the study.

After obtaining support and approval at board level for each health board region, purposeful sampling was used to select and approach a mixture of small, medium and large general practices. This included remote and rural practices that are often excluded from traditional research participation. Practices were approached using a range of methods including emails with promotional material, telephone calls and invited presentations at practice meetings. A database was maintained to track responses from invited practices and where possible reasons for declining participation were recorded.

Later in the pilot, in preparation for larger-scale region-wide practice recruitment, we developed a training program for primary care pharmacy staff. This was delivered in-person or via regional online study and training events, and provided education and training on prescribing switch implementation, as well as facilitating practice recruitment.

### Identification of practice study population

Following recruitment of a practice, suitable patients were identified using a bespoke electronic search tool developed during the pilot, deployed within the practice electronic records system. We developed the tool to work flexibly on the two dominant practice information systems used in Scotland, EMIS (www.emishealth.com) and VISION (https://info.visionhealth.co.uk/gp-solution). This evolved iteratively during the pilot, becoming progressively more effective and efficient at correctly identifying the appropriate study population. All patients who met the inclusion/exclusion criteria were included in the study. The inclusion and exclusion criteria are detailed in Table 1.

**Table 1:** Patient search criteria.

| Inclusion Criteria | Exclusion Criteria |
| --- | --- |
| Documented diagnosis of hypertension (recorded on practice hypertension register) | Documented history of an adverse drug reaction to either medication |
| Currently receiving repeat prescriptions of bendroflumethiazide tablets or indapamide tablets | History of having been prescribed both thiazide-like diuretics at different times, indicating a potential clinical reason for not being able to take one or the other |
| Aged 18 and over | Other clinical or non-clinical indication for not switching medication (applied manually, see below) |

A provisional list of patients produced by the search was further reviewed by a GP or suitable member of practice staff based on their knowledge of the patients. This review could result in further exclusions. The resulting list then formed an approved practice study population.

### Randomisation

Randomisation was 1:1, block-balanced by practice list size, using a computerised randomisation algorithm applied after identification of the final practice study population. Where required by the randomised policy repeat prescriptions for patients in the practice study population were then switched on the practice patient medication system as follows:

1. Bendroflumethiazide 2.5mg, 5mg or indapamide 1.5mg (slow release) was changed to indapamide (standard release) 2.5mg
2. Indapamide 2.5mg, 1.25mg, 1.5mg (slow release), or bendroflumethiazide 5mg was changed to bendroflumethiazide 2.5mg.

The selected standard dosages are in line with most national and regional formularies for thiazides use in hypertension (20).

The study pharmacist facilitated any prescription changes required within the practice using the usual process employed by the practice for any mass prescription changes.

### Comparing outcomes between the two prescribing policies

We plan that the primary and secondary outcomes will be obtained through anonymised routinely acquired national health data sets using eDRIS and GP practice data using trusted third-party services provider, Albasoft. Permissions have been obtained from the Public Benefit and Privacy Panel to process these data.

#### Primary Outcome

- Fatal or non-fatal myocardial infarction, coronary revascularisation, fatal or non-fatal stroke, fatal or nonfatal heart failure or vascular death.

#### Secondary Outcomes

- Individual components of the primary outcome
- All-cause mortality
- Metabolic complications (hypokalaemia and hyponatraemia)
- New diabetes mellitus diagnoses

### Informing patients about the study

Immediately following randomisation of the practice prescribing policy, all patients in the study population were informed of the research study and potential medication change. This was communicated by letter and distributed via an NHS approved third party mailing company (Docmail). The letter was printed on standard practice headed paper specific to each practice, and briefly explained why the study was being conducted and advised that the patient’s repeat prescriptions for thiazide-type medication might be switched in line with the newly assigned practice prescribing policy. The letter directed patients who may have any questions, concerns, or objections to visit a study-specific website (www.memoresearch.com/evidence) or to contact the study team directly.

After the prescribing policy implementation in each participating practice, information packs were provided to be dispatched by the practice to all local community pharmacies, informing them of the study and the randomisation. This pack also contained a copy of the patient letter, and an information poster to display if they wished.

### Tracking study workload

In our efforts to reduce to a minimum the impact of our study on general practice workload we directed any queries to the study team. This required a means of tracking all contacts whether via telephone, email or the EVIDENCE website. We therefore created a database to enable us to record and collate all contacts received from patients and staff. This database facilitated the assimilation of feedback, allowing for iterative improvements in the study design and ensured that all patients were provided with the information they required promptly. If a patient preferred to opt-out of the switch and remain on their current medication, the study team contacted the surgery to reverse the switch for them before their next prescription issue; thus, avoiding any disruption to their medication supply.

## Results

### Practice Recruitment

In prior consultations with stakeholders, we identified that the principal challenge to be overcome in recruiting practices was concern about increased workload. In response to this we developed the study methods that allowed for minimising disruption to practice workflows to negligible levels. Furthermore, the study processes are designed to be implemented by study pharmacists or by locality pharmacists trained by the study pharmacist and working temporarily under the aegis of the study team. Currently 29 pharmacists and pharmacy technicians across Scotland are delegates that support the implementation of the EVIDENCE study. in addition to the core study team. We found that endorsement of the study at health board-level greatly facilitated acceptability by practices by minimising concerns about potential impact on prescribing budgets, staff time, and possible penalties for regional formulary non-adherence.

Figure 1 provides an overview of practice recruitment and the study population. Prior to Covid-19, out of 77 practices approached in 5 Scottish health board regions (Highlands and Islands, Grampian, Fife, Dumfries and Galloway and Tayside), 33 (43%) agreed to participate, 16 (21%) declined, and 28 (36%) did not respond. Despite attempts to assure practices that every effort would be made to minimise the workload consequence of taking part in the study, the dominant reasons for declining to participate were the practice being “too busy” or “short of staff”. No practices approached gave disagreement with the premise of the study as a reason not to take part.

**Figure 1.**
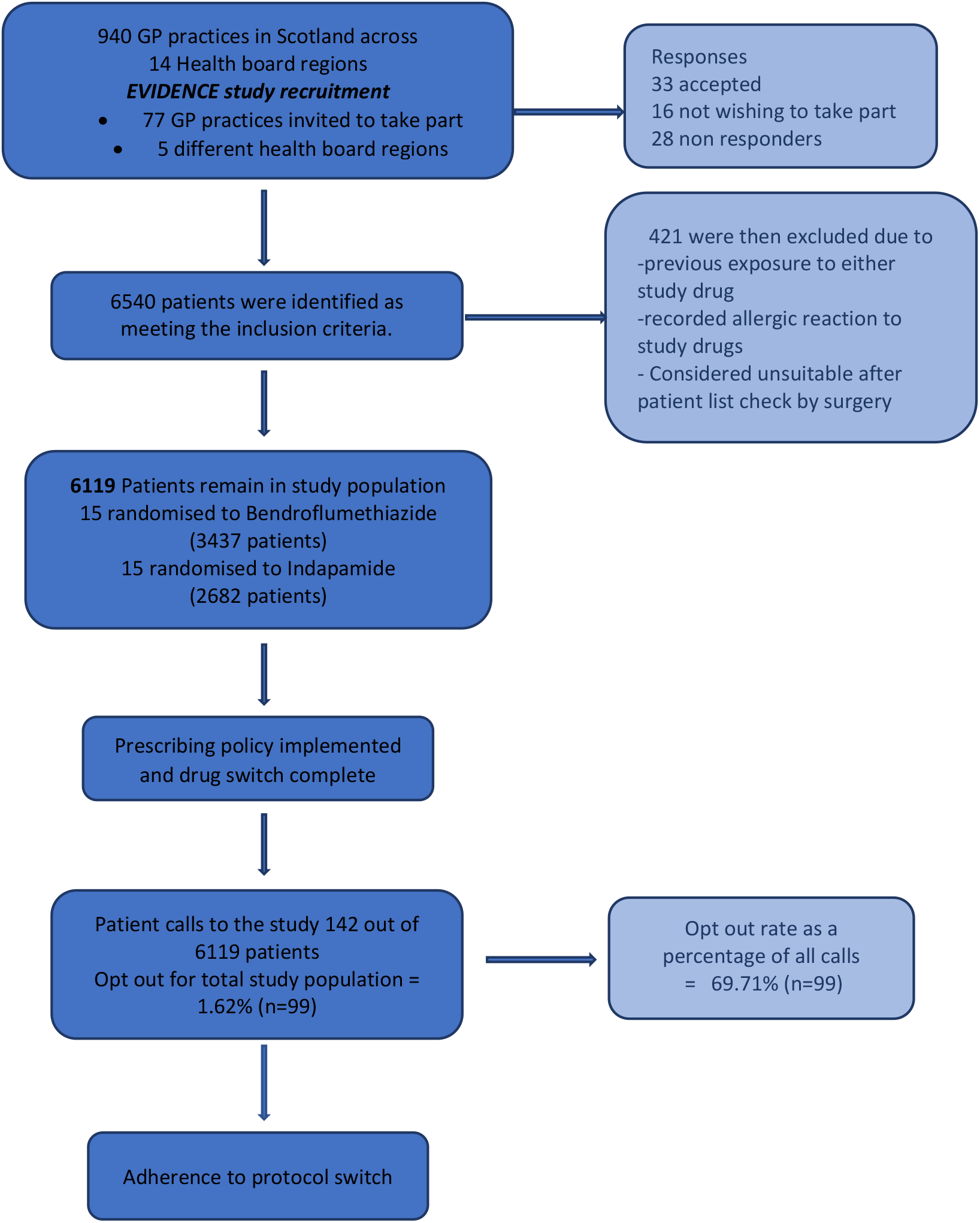
Consort style flow chart showing results of practice recruitment to pilot study and resulting study population including patient opt out percentage.

We found that a key incentive for practices to participate was to support training and research in primary care pharmacy, particularly in improving the efficiency of routine drug switching exercises, for example, due to changes in regional prescribing policy. We tailored our recruitment approach to emphasise the training and efficiency opportunities of taking part and the progression of an embedded research culture. As the pilot study progressed, and the study processes became more streamlined, we found that the perceived acceptability by practices improved.

In the pilot phase of this study 30 practices were recruited and had the randomised policy implemented. This included 3 (10%) practices classified as remote and rural by the NHS Scotland Information Statistics Division (ISD) (21). Table 2 provides a breakdown of practice recruitment by NHS health board region.

**Table 2:** Practice recruitment.

| ***Health board*** | ***Number of practices Contacted*** | ***Number of Positive responses*** | ***Number of Negative responses*** | ***No response*** |
| --- | --- | --- | --- | --- |
| Fife | 24 | 5 | 7 | 12 |
| Tayside | 39 | 22 | 7 | 10 |
| D&G* | 10 | 3 | 2 | 5 |
| Grampian | 3 | 2 | 0 | 1 |
| Highland | 1 | 1 | 0 | 0 |
| ***Total*** | ***77*** | ***33 (43% of total contacted)*** | ***16 (21% of total contacted)*** | ***28 (36% of total contacted)*** |

\*Dumfries & Galloway

### Study population and randomisation

The 30 completed practices had a total registered population of 187,106 patients with a mean list size of 6237 patients (range 1808 to 12778). After refinements to the study tool, we included a search to identify patients who were recorded on the hypertension register. The mean proportion of patients with a diagnosis of hypertension in those practices was 15.2% (range 9.0 to 19.8%) of the list size. After applying the exclusion criteria to the hypertension register patients, we found 19.0% (13.1-27.2%) of these remained leaving a total study population of 6119, or approximately 208 patients per practice (range 44 to 539). By the end of the pilot, the search and repeat prescription switching processes in one practice took on average 12 working hours (1.5 working days) for a trained pharmacist to undertake.

After randomisation across 30 practices, 15 practices (3437 patients) were allocated to the bendroflumethiazide prescribing policy and 15 practices (2682 patients) to the indapamide policy.

### Patient acceptability

In total, 142 patients (2.3% of the study population of 6119 patients) contacted the study team after receiving the notification letter; a mean of 5.1 patient calls per practice (range 0 to 15). Table 3 shows a breakdown of reasons for patient calls received by the study team. Because patients could have more than one reason for contacting the study team, the number of reasons (160) exceeds the number of individual patients.

**Table 3:** Results from patient call tracker.

| ***Reason for contact \**** | **n** | **% of contacts** |
| --- | --- | --- |
| Request for more information | 53 | 37.3 |
| Concern about the risk of adverse event | 32 | 22.5 |
| Adverse event to a non-study drug | 4 | 2.8 |
| Opt out of switch due to general ill-health | 6 | 4.2 |
| Opt out of switch for a non-specific reason | 55 | 38.7 |
| General feedback on the letter | 6 | 4.2 |
| Concern about ethical aspects of the study | 4 | 2.8 |
| ***Outcome of patient contact*** |  |  |
| Prepared to switch | 43 | 30.3 |
| Not prepared to switch | 99 | 69.7 |
| **Total number of patients** | **142** |  |

\* Patients may have made contact for more than one reason

The most common reasons for contact were requesting more information or not wishing to be switched (37.3% and 38.7% respectively of total number of patient calls). Another significant proportion of calls were due to concerns about potential side effects of the new medication. The outcome of 99 patient contacts with the study team was the patient wishing to refuse a drug switch; this represented only 1.62% of the total study population. Only a tiny proportion of patients expressed any unsolicited concern over the ethics of the study (2.8% of all patient contacts with the study team and less than 0.01% of the study population). The feedback from these patients was extremely helpful in improving the content of the communication letter and the manner and timing of prescribing changes. As these improvements were progressively implemented the number of patient calls per practice reduced.

More patient calls were received from practices randomised to the indapamide policy n=85, (3.17% of the indapamide study population), than to the bendroflumethiazide policy n=57 (1.66% of the bendroflumethiazide study population). Though the practices were not required to record the number of patients contacting them independently, feedback from the practice staff has indicated that this was minimal.

### Adherence

Analysis of aggregate practice-level prescribing data (www.opendata.nhs.scot/dataset/prescriptions-in-the-community) showed that, amongst practices randomised to bendroflumethiazide, the proportion of indapamide prescribing before policy randomisation was about 20%; dropping to 9% afterwards. For practices randomised to indapamide, the proportion of prescriptions for indapamide prior was 16%; increasing to 82% (Figure 2). This data also shows that there has been little evidence of switching back with time, consistent with the very low numbers of individuals who initially refused a medication change. Note that the residual prescribing of non-randomized policy drug in the above data could also have been due to these being prescribed for indications other than hypertension.

**Figure 2.**
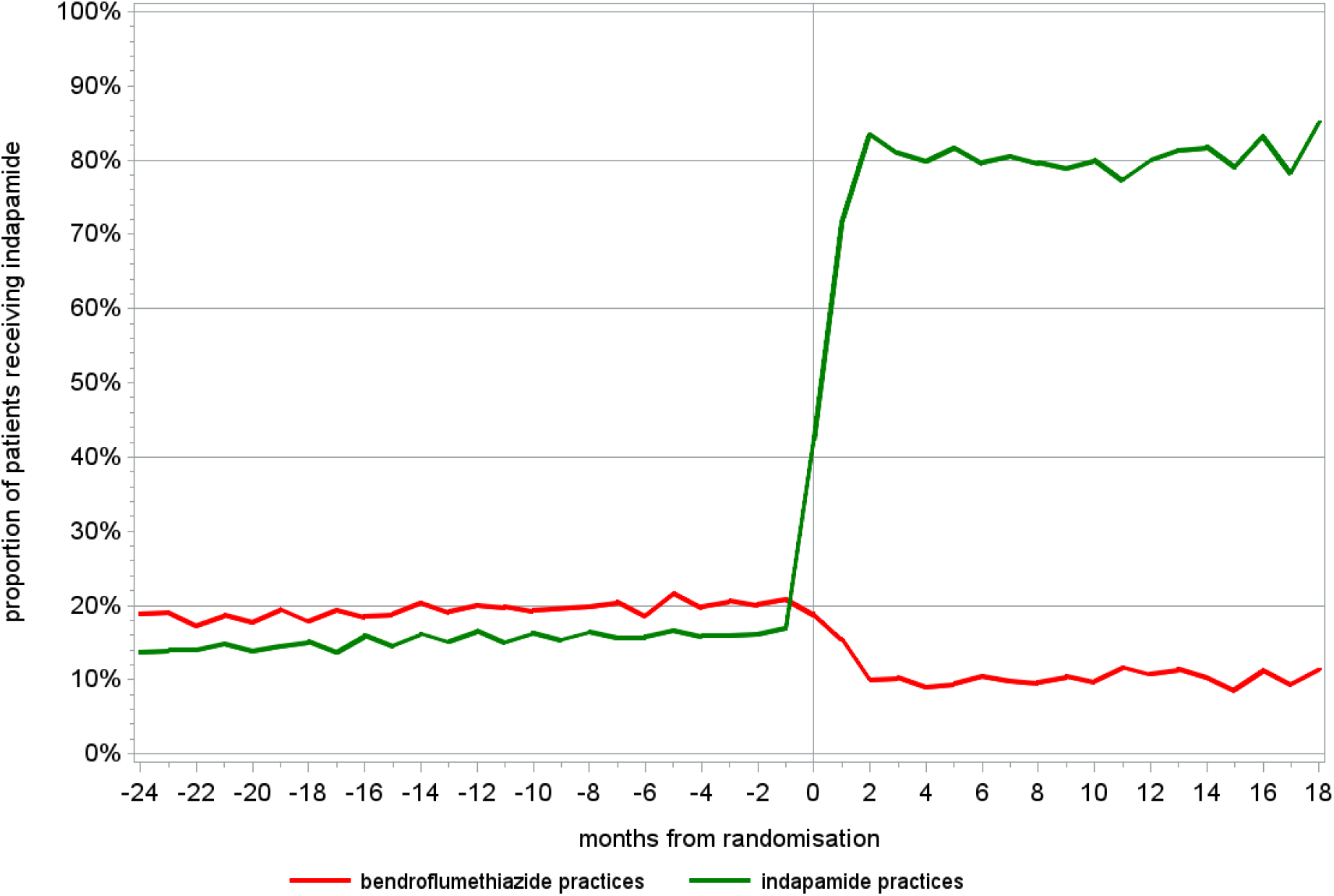
A plot showing practice adherence following prescription switches over time to the randomised policy for practices randomised to bendroflumethiazide and indapamide.

## Discussion

This pilot study has demonstrated the feasibility and acceptability of the EVIDENCE study design. By randomising only thirty practices to a prescribing policy of either indapamide or bendroflumethiazide, we rapidly generated a study population of over 6000 individuals at a relatively low cost. The pilot has also informed improvements in methodological approaches to increase acceptability, efficiency, and scalability for higher practice recruitment rates in future. Importantly, we received only trivial amounts of negative feedback from patients and healthcare practitioners involved, with the majority of this being during the early stages of the pilot. Main concerns raised by practice staff included differences in cost of drugs and adherence to regional formulary, along with lack of staffing resources to implement the study. This feedback was invaluable in improving the trial protocol.

### Practice Recruitment

Scoping work prior to the pilot indicated that resource pressures in primary care would challenge recruitment and implementation for any type of practice-based research. This was reinforced by feedback gathered from practice staff during the recruitment process which showed that the most frequent reasons given for not wishing to take part were challenges with staffing levels and general workload. This greatly influenced the EVIDENCE study design which aims to produce minimal workload for practice staff and a streamlined processes for study staff. Neither practice staff nor the wide range of other stakeholders approached during the pilot expressed concern over the concept of randomisation of prescribing policy and subsequent prescription switching. Neither did any healthcare professional express concern over prescribing policy randomisation in principle. There was an overall acceptance and intrinsic understanding of the need for this type of research within the background of regular drug switching within the NHS, often done for cost difference with the assumption of equivalent effectiveness.

Although arranging visits to individual practices was useful in engaging and obtaining feedback from GPs and other practice staff, we found this approach time consuming and resource intensive. An email plus direct telephone approach was more time-efficient and equally effective. During the pilot, we further evolved the recruitment strategy by obtaining endorsement of the study at a regional level. This high-level support significantly reduced individual practice concerns about taking part. NHS board endorsement also facilitated collective recruitment and training of primary care pharmacy staff at regional training days. This has been further adapted throughout COVID-19 to allow for remote training methods and study support, proving a useful resource during current circumstances and in the future. Three health boards (Lothian, Fife and Dumfries and Galloway) have since agreed to implement the full study in this way. With the experience and knowledge gained by undertaking the pilot we have since been able to scale recruitment with currently over 100 practices across Scotland recruited comprising over 12,700 patients.

### Patient Acceptability

In a previous public opinion study, it was found that the public in Scotland is broadly supportive of the concept of randomised policy design studies of medicines. At the same time, there was a spread of opinion among GPs (18). Consistent with this, the number of patients refusing a drug switch was very low, and aggregate prescribing data has confirmed that trivial numbers of individuals switched back. As might be expected, a higher number of patient contact calls were received from practices randomised to indapamide, where more drug switches were required, and therefore more patients were being affected by the change in prescribing policy. We have continually sought feedback from patients, healthcare professionals, public involvement groups, the research ethics committee, and other stakeholders throughout this pilot. Their feedback has allowed us to make incremental improvements to the study methods and patient communications; this has included: -

- making the choice to refuse a drug switch much clearer and more prominent to patients.
- a link to the study website has been added to the patient letter to allow a greater amount and detail of information to be presented.
- information posters about the study are given to participating practices and community pharmacies to display either physically or online.

### Study implementation

Pressures of clinical workload and limited resources available are recognised barriers to participation in primary care research. This issue was at the forefront of developing the methodology and recognition of practice acceptability for the study. The pilot has allowed the refinement of an electronic search tool to identify a study population efficiently, reducing the need for manual medical history reviews. This, along with other improvements, allowed rapid large-scale operations to be carried out in an efficient and effective manner. For example, during the initial few practices, for a single practice it could take several weeks to move from practice recruitment to completion of switching. Multiple factors influenced this process, such as physical space in practice to allow the work to be carried out, insufficiently specific search criteria for patient identification, and delays within the surgery in authorising of the patient list. This pilot has allowed many of these barriers to be addressed. The time required to implement a policy switch has also been greatly improved, for example, later in the process, a practice with a list size of over 10,000 patients was processed within a single working day.

### Integration with existing NHS staff and multidisciplinary working

By engaging with NHS staff and incorporating existing NHS prescribing processes, we have achieved the potential to rapidly expand the study in regions that are too distant for research centre staff to travel to easily. Also, we have contributed to the development of infrastructure that supports pharmacy practice development, education, research and electronic sharing of information, an ideal set out in the Prescription for Excellence government strategy (22). Pharmacotherapy services, as part of the new Scottish GP contract, also reinforces the role of pharmacy in prescribing policy implementation. Our pragmatic study design is aligned with these national objectives as chronic disease management is often under the remit of the practice pharmacist. Pharmacy staff at both health board and practice level have engaged with the study as a way of enhancing their skills and developing a research ready workforce.

## Conclusion

By attending to some well-established challenges to clinical trials recruitment, this pilot has effectively demonstrated that significant expansion of this study is not only feasible but will provide a prescribing policy framework that can be adapted to many therapeutic areas. By incorporating randomisation to the routine practice of prescribing policy changes we aim to enable formal evaluation of comparative effectiveness and produce research outcomes efficiently with minimal disruption to patients and routine clinical practice and at low cost.

Engagement from key stakeholders has assisted recruitment of practices and aided policy implementation through regional health board pharmacy staff. In addition, remote access to GP systems has become more available and we have utilised this technology to minimise practice footfall and inconvenience, essential during the recent pandemic.

Following the positive results of the pilot work described above we have continued to progress the EVIDENCE study and as of July 2021 102 GP practices (6 Scottish Health Boards) are currently recruited, 27 regional pharmacy delegates are assigned to implement the study and approximately 12,800 patients have been subject to the randomised policy design. The successful pilot and subsequent expansion demonstrate the effectiveness of this multidisciplinary and pragmatic trial approach to health services research and the willingness to engage in delivering evidence-based medicine despite challenges of covid-19 restrictions.

## Data Availability

Not applicable but any data refered to can be requested from the author

## Abbreviations

NHS: National Health Service
WHO: World Health Organization

## Declarations

### Consent for publication

Not applicable.

### Availability of data and materials

Not applicable.

## Authors contributions

TM and IM devised the pilot study methodology. AF, AD, AR and LM ran the pilot study. RF provided statistical analysis. AF drafted the initial manuscript, and all authors contributed to developing the manuscript. AF edited and finalised the manuscript. All authors read and approved the final manuscript.

## Acknowledgements

We acknowledge the contributions of the NHS Scotland general practice and trust staff, including doctors, pharmacists, practice managers and administrators, who advised on appropriate methods, as well as the patients who have taken the time to provide feedback on their experience. We would also like to thank the health board pharmacy leads that helped to deliver this study and continue to support the development of this important research.

## REFERENCES

1. Gagne JJ, Choudhry NK. How Many “Me-Too” Drugs Is Too Many? JAMA. 2011;305(7):711–2.

2. Cipriani A, Ioannidis JPA, Rothwell PM, Glasziou P, Li T, Hernandez AF, et al. Generating comparative evidence on new drugs and devices after approval. The Lancet. 2020;395(10228):998–1010.

3. Naci H, Salcher-Konrad M, Kesselheim AS, Wieseler B, Rochaix L, Redberg RF, et al. Generating comparative evidence on new drugs and devices before approval. The Lancet. 2020;395(10228):986–97.

4. Guthrie B, Boyd CM. Clinical Guidelines in the Context of Aging and Multimorbidity. Public Policy & Aging Report. 2018;28(4):143–9.

5. Saunders C, Byrne CD, Guthrie B, Lindsay RS, McKnight JA, Philip S, et al. External validity of randomized controlled trials of glycaemic control and vascular disease: how representative are participants? Diabetic Medicine. 2013;30(3):300–8.

6. Ware JH, Hamel MB. Pragmatic Trials — Guides to Better Patient Care? New England Journal of Medicine. 2011;364(18):1685–7.

7. Treweek S, Lockhart P, Pitkethly M, Cook JA, Kjeldstrom M, Johansen M, et al. Methods to improve recruitment to randomised controlled trials: Cochrane systematic review and meta-analysis. BMJ open. 2013;3(2).

8. National Institute for Health Research. Improving inclusion of under-served groups in clinical research: Guidance from INCLUDE project 2020 [Available from: https://www.nihr.ac.uk/documents/improving-inclusion-of-under-served-groups-in-clinical-research-guidance-from-include-project/25435.

9. England. PH. Health matters: combating high blood pressure. 2017 [Available from: https://www.gov.uk/government/publications/health-matters-combating-high-blood-pressure/health-matters-combating-high-blood-pressure.

10. Lawes CM, Vander Hoorn S, Rodgers A. Global burden of blood-pressure-related disease, 2001. Lancet (London, England). 2008;371(9623):1513–8.

11. Moser M, Feig PU. Fifty years of thiazide diuretic therapy for hypertension. Archives of internal medicine. 2009;169(20):1851–6.

12. National Institute for Health and Care Excellence. NICE Update of clinical guidelines 18 and 34 Hypertension. The clinical management of primary hypertension in adults Clinical Guideline 127 Methods, evidence, and recommendations August 2011.

13. National Institute for Health and Care Excellence. Hypertension in adults: diagnosis and management. NICE guideline [NG136] Published date 28 August 2019

14. Brown MJ, Cruickshank JK, Macdonald TM. Navigating the shoals in hypertension: discovery and guidance. BMJ (Clinical research ed). 2012;344:d8218.

15. Public Health Scotland. Community Dispensing [Available from: https://www.isdscotland.org/Health-Topics/Prescribing-and-Medicines/Community-Dispensing/Dispenser-Remuneration/.

16. Macfarlane TV, Pigazzani F, Flynn RWV, MacDonald TM. The effect of indapamide vs. bendroflumethiazide for primary hypertension: a systematic review. British journal of clinical pharmacology. 2019;85(2):285–303.

17. Rogers A, Craig G, Flynn A, Mackenzie I, MacDonald T, Doney A. Cluster randomised trials of prescribing policy: an ethical approach to generating drug safety evidence? A discussion of the ethical application of a new research method. Trials. 2020;21(1):477.

18. Mackenzie IS, Wei L, Paterson KR, Macdonald TM. Cluster randomized trials of prescription medicines or prescribing policy: public and general practitioner opinions in Scotland. British journal of clinical pharmacology. 2012;74(2):354–61.

19. Rogers A, Flynn A, Mackenzie IS, McConnachie L, Barr R, Flynn RW, et al. Evaluating Diuretics in Normal Care (EVIDENCE): Protocol of a cluster randomised controlled equivalence trial of prescribing policy to compare the effectiveness of thiazide-type diuretics in hypertension. medRxiv. 2020:2020.12.23.20248767.

20. Joint Formulary Committee. Joint Formulary Committee. British National Formulary: London: BMJ Group and Pharmaceutical Press [Available from: https://bnf.nice.org.uk/treatment-summary/diuretics.html.

21. National Services Scotland. General Practice – GP workforce and practice list sizes 2005-2015.; 2015.

22. The Scottish Government. Prescription for Excellence. A Vision and Action Plan for the right pharmaceutical care through integrated partnerships and innovation.; 2013.

